# Distribution of Smoking and Hypertension: A Cross-Sectional Study of Bangladeshi Beverage Company Staff

**DOI:** 10.1101/2024.11.14.24317335

**Authors:** Abu Sayeed Md. Saleh, Tilat Shaharin, Afnan Shafiullah, S M Saidur Rahman Mashreky

**Affiliations:** Department of Public Health, North South University.

**Keywords:** Noncommunicable diseases (NCDs), Premature mortality, Cardiovascular diseases, Hypertension, Risk factors, Unhealthy diet, Physical inactivity, Obesity, Nicotine, Coronary heart disease, Aneurysms, Peripheral arterial disease, Buerger’s disease, Cigarette smoking

## Abstract

Non-communicable diseases, especially hypertension, are disproportionately problematic in low- and middle-income countries such as Bangladesh, where they have become one of the leading causes of death among its population. The present study compares both the prevalence of hypertension and smoking in employees of Bangladeshi beverage industries to that of the general population. By capitalizing on data from various sources, this research evinces that the prevalence of the preceding issues is disturbingly high, particularly among certain demographics. This, despite the efforts the National Heart Foundation put in place to address the challenges in hypertension screening and management, reflects the need for healthcare delivery. The connection between smoking with health in this sense is confirmed; thus, it will be of crucial necessity to understand the link between these threats, especially within the beverage industry. This study has, therefore, tried to shed light on the nexus, to inform targeted interventions aimed at promoting healthier work environments for the ultimate reduction of NCD-related deaths within the country.

## Introduction

NCDs are a silent killer in the sphere of global health and tend to affect more people from low- and middle-income groups of countries. In 2016, NCDs accounted for 71% of the total 57 million deaths recorded worldwide; this includes 17.9 million deaths due to cardiovascular diseases alone. High blood pressure, generally known as hypertension, is one of the major players in this scenario, wherein unhealthy diets, insufficient levels of physical activity, and tobacco and alcohol consumption play a major role.

Tobacco smoke is a toxic cocktail of chemicals, including nicotine, which has been shown to cause a range of cardiovascular problems, including coronary heart disease, strokes, and peripheral arterial diseases [1,2]. Its pernicious effects do not stop at the heart but extend to a range of cancers and respiratory illnesses [3]. Not surprisingly, both smoking and hypertension are recognized as cardinal risk factors for cardiovascular disease [4,5,6], contributing in major ways to preventable morbidity and mortality worldwide [1].

While indeed the global trend is toward living longer, developing countries like Bangladesh continue to grapple with many and varied complex issues. This rapidity of this aging process outstrips all other measures of development in society and the economy, and there has also been a disturbing decline in physical activity and a disturbing increase in the consumption of alcohol and tobacco. Bangladesh has a dual burden of having one of the highest male smokers-33.2% among adults aged 18+ years-nationwide- and one of the highest hypertension rates in the world −11.2 % among adult women and as high as 37.4% among adult men [1,7]. GATS 2017 depicts a grim scenario where tobacco products, mainly in the form of cigarettes, are being consumed by a sizable section of the population of Bangladesh.

In this regard, focused studies targeting the beverage industry employees in Bangladesh become an urgent need. The prevalence of hypertension and smoking habits among employees of such industries and their comparison with the general population will be carried out in the present study. Addressing these knowledge gaps by proposing the conduct of appropriate interventions, the study hopes to explore not only the prevalence of hypertension and its determinants but also the awareness of the potential of lifestyle modifications. This will, it is hoped, bring a better understanding of the association between hypertension and smoking habits among the beverage industries in Bangladesh and help establish healthier workplaces for all employees.

Prevalence estimates range from 16% to as high as 40% in Bangladeshi adults. It is highest among the elderly, reaching as high as 40% in that age bracket. This is fuelled by socioeconomic status, urbanization, and therefore, also obesity. In addition, studies have highlighted marked differences in the prevalence of undiagnosed hypertension across different demographic groups in Bangladesh.

Hypertension has been a challenge, with continuous awareness, treatment, and control issues despite active efforts in Bangladesh. The National Heart Foundation takes part in global hypertension screening activities, though there are deficiencies regarding routine clinical screening practices [8,15]. This indicates that improvement in strategies to deal with hypertension needs to be systematically developed throughout Bangladesh [16,15].

Smoking prevalence is another important public health issue in Bangladesh: It was very disheartening when the 2017 GATS showed that 40% of males and a very disconcerting 25.2% of females use tobacco [7]. While there has been an overall decline in tobacco use, male smoking remains very high at 37% [7]. However, in subgroups, such as ready-made garment workers, smoking prevalence is even higher, with reported figures reaching a high of 86.2% among rural men [17, 18].

Smoking has been identified as a risk for COPD, chronic back pain, and ischemic stroke [19,20,21]. There is even regional variation: hypertension was found to be more prevalent in coastal areas than elsewhere, which further reinforces the need for geographical equity to be considered [22]. Poor mental health in Bangladeshi university students is associated with smoking behavior [23].

The high prevalence of smoking in certain sections of the population in Bangladesh, in conjunction with the increasing trend in hypertension prevalence observed, reinforces suspicion of a link between these two health hazards. Only further targeted research can establish a comprehensive understanding of this complex relationship.

## Materials and Methods

### Data Collection Tools

- WHO STEPS Questionnaire
- Sphygmomanometer
- Stethoscope

### Data Collection Methods

Study design and Target population: This is a descriptive type of cross-sectional study. This study was conducted among Employees of a Bangladeshi beverage company in the district of Dhaka. Both management and non-management staff were selected by convenient sampling and interviewed for this study. The age range of the study participants was from 20 to 60 years. BP was measured using a mercury sphygmomanometer whose cuff size was 12 cm wide and 22 cm long. The SBP and DBP were measured by a trained nurse. Two measurements of BP were made with a 10-minute interval between the two measurements. The participants during these measurements would sit with their left arm at the heart level supported with their palm facing upwards. The mean of the two readings was taken as the participant’s BP. The survey was performed using the WHO STEPwise approach to surveillance of noncommunicable disease (STEPS) with other validated questionnaires and other technical publications by taking expert advice from various sources. A pilot study was done to stabilize the questionnaire according to the current research environment and also to increase the reliability and validity of the study. The socio-demographic variables like age, sex, smoking habits, and the risk factors for cardiovascular diseases were obtained by using the questionnaire.

### Measurements

The STEPS survey included measures on demographic characteristics; behavioral risk factors of smoking, fruit and vegetable consumption, and physical activity, as well as physical characteristics of weight, height, and blood pressure. All measurements were performed in conformation with the WHO STEPS protocols.

Hypertension was defined by the presence of at least one of three conditions: (a) SBP⩾140 mm Hg; (b) DBP⩾ 90 mm Hg; or (c) current use of anti-hypertensive drugs. Smoking status was classified as ‘Yes’ or ‘No’.

Height was measured to a precision of 1 cm using a stadiometer on each participant. Participant weight was measured in light clothing without shoes using a precision scale of 100 g. Body mass index was defined as weight in kilograms divided by height in meters squared. Each indicator was measured three times on each subject. Then the mean was taken for analysis. The highest education attained by each respondent was categorized into Secondary Education, Higher Secondary Education, and Graduate.

All the participants provided written informed consent. This study was reviewed and approved by the Research Ethics Committee of North South University, Dhaka, Bangladesh under the approval reference number #2024/OR-NSU/IRB/0311. The approval was granted on 2-April-2024. All procedures performed in the study involving human participants were in accordance with the ethical standards of the institutional and national research committee and with the 1964 Helsinki Declaration and its later amendments or comparable ethical standards.

### Statistical analysis

This section describes the statistical methods used to analyze data from the study. Descriptive statistics were utilized in the process of characterization of participants, while chi-square tests were used in identifying risk factors.

Descriptive statistics was used to summarize data on both categorical and continuous variables. The frequency and percentage were computed for the categorical variables to display a distribution of responses in each category. In respect of the continuous variables, the mean and standard deviation were computed to reflect the average value and spread of the points, respectively.

Chi-square tests were then performed to assess the associations of group membership-hypertension and smoking status with all categorical variables. The χ2 was reported in each test with its p-value. the association between the two variables was considered statistically significant If the p-value was found to be less than 0.05.

Statistical analyses were performed using the IBM SPSS Statistics for Windows [Version 27.0]. The two-tailed P ≤ 0.05 was taken as the level of statistical significance.

### Results and Findings

**Table 1:**
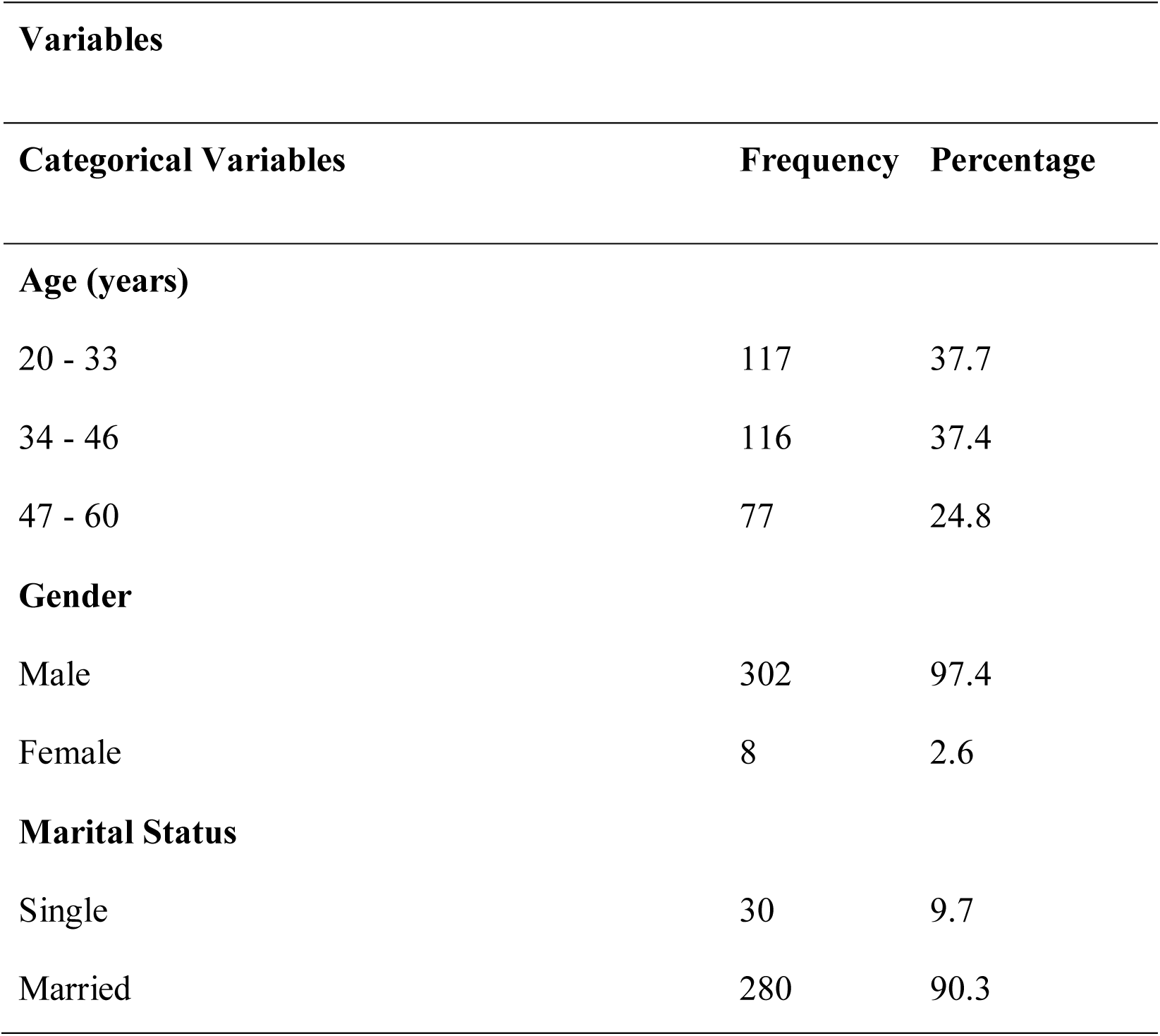
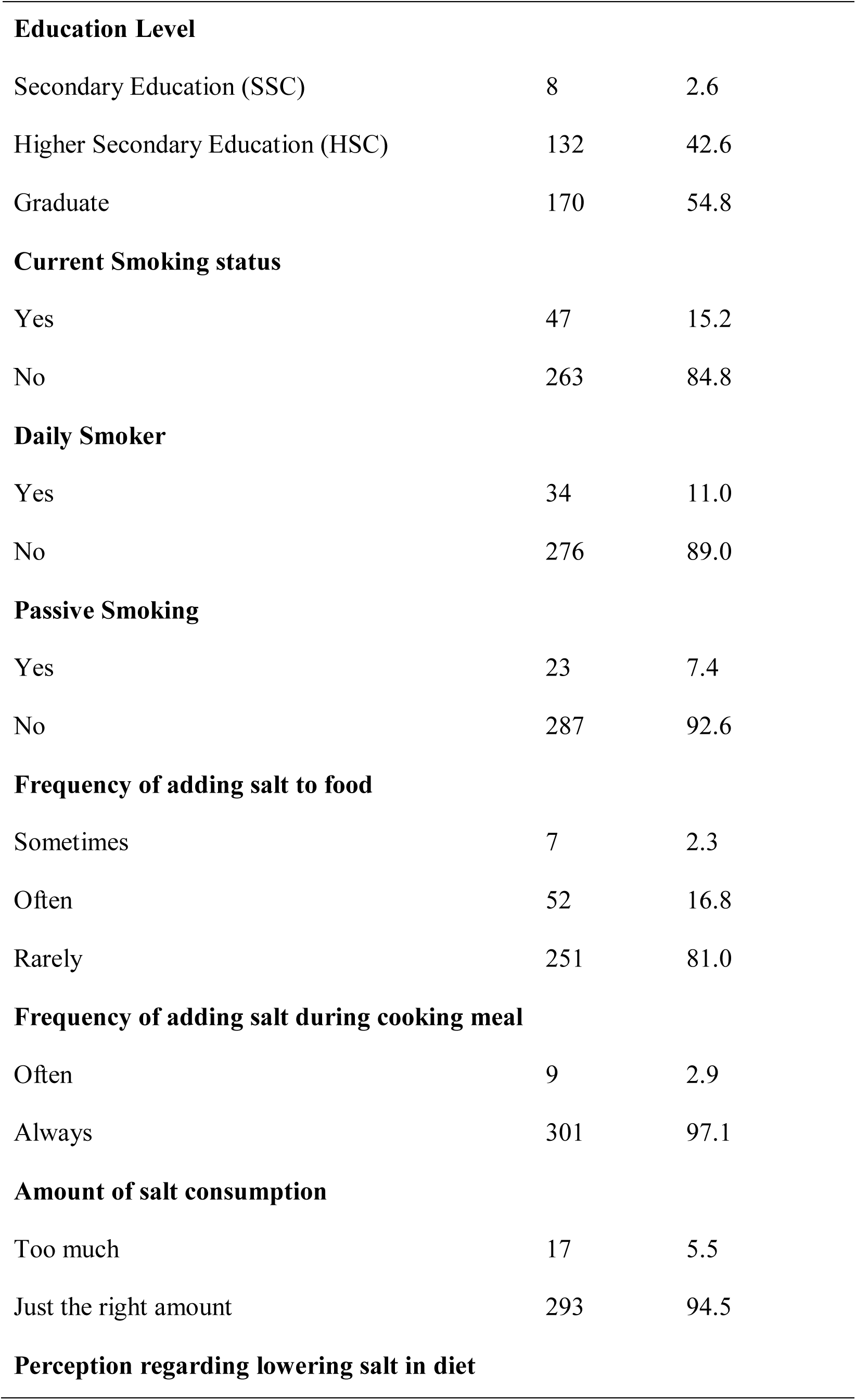
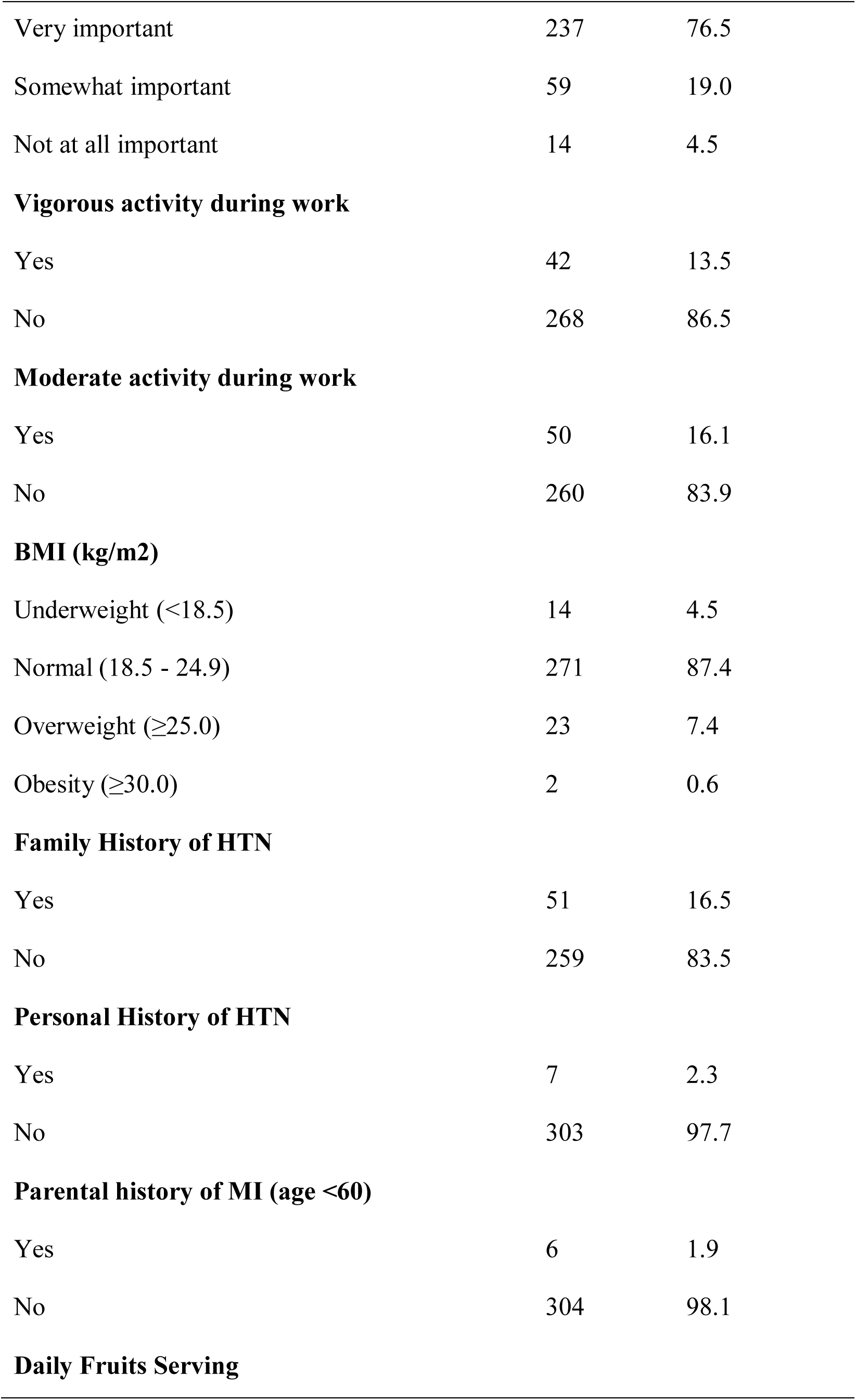
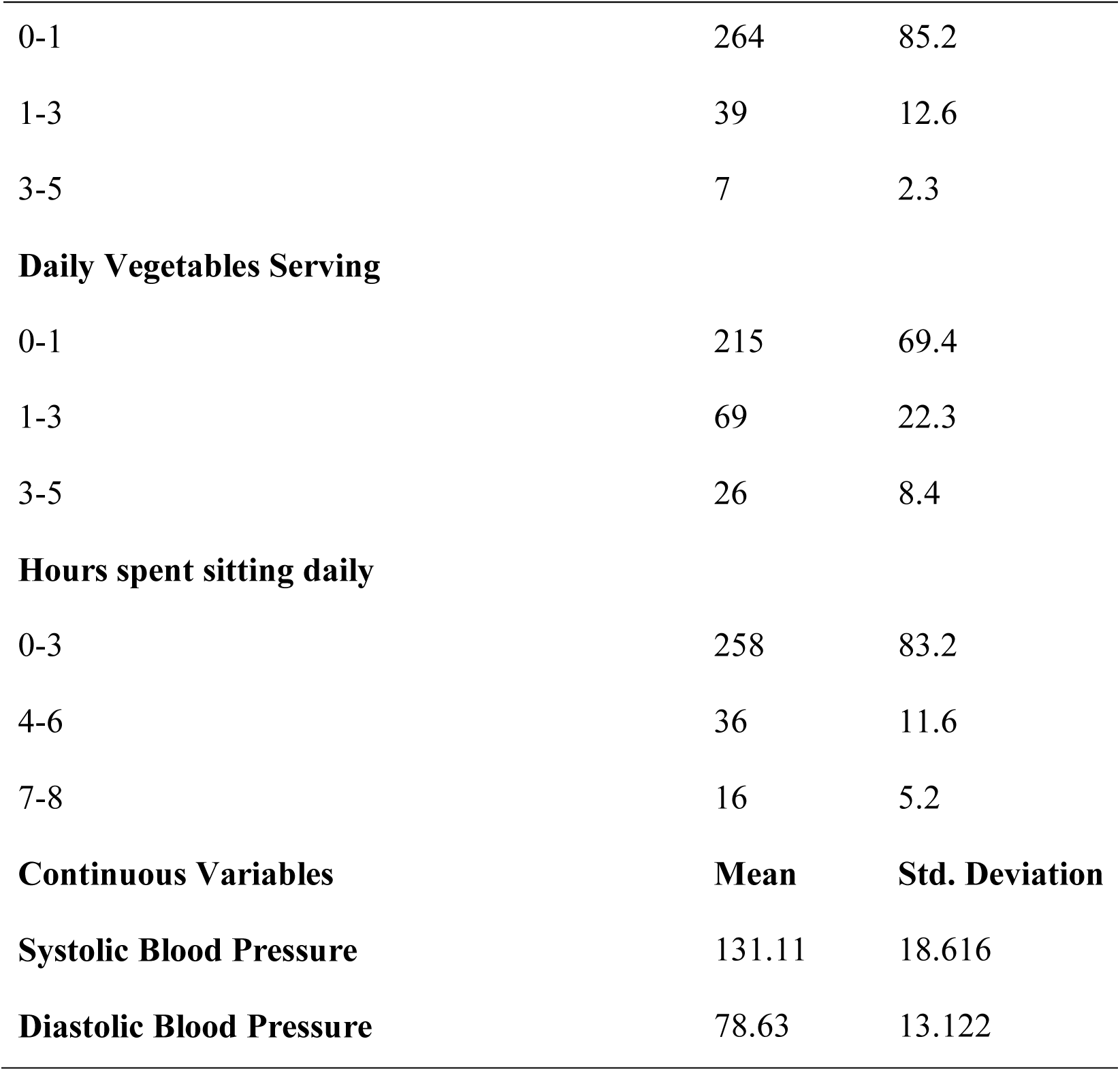
Descriptive statistics.

| **Variables** |  |  |
| --- | --- | --- |
| **Categorical Variables** | **Frequency** | **Percentage** |
| **Age (years)** |  |  |
| 20 - 33 | 117 | 37.7 |
| 34 - 46 | 116 | 37.4 |
| 47 - 60 | 77 | 24.8 |
| **Gender** |  |  |
| Male | 302 | 97.4 |
| Female | 8 | 2.6 |
| **Marital Status** |  |  |
| Single | 30 | 9.7 |
| Married | 280 | 90.3 |
| **Education Level** |  |  |
| Secondary Education (SSC) | 8 | 2.6 |
| Higher Secondary Education (HSC) | 132 | 42.6 |
| Graduate | 170 | 54.8 |
| **Current Smoking status** |  |  |
| Yes | 47 | 15.2 |
| No | 263 | 84.8 |
| **Daily Smoker** |  |  |
| Yes | 34 | 11.0 |
| No | 276 | 89.0 |
| **Passive Smoking** |  |  |
| Yes | 23 | 7.4 |
| No | 287 | 92.6 |
| **Frequency of adding salt to food** |  |  |
| Sometimes | 7 | 2.3 |
| Often | 52 | 16.8 |
| Rarely | 251 | 81.0 |
| **Frequency of adding salt during cooking meal** |  |  |
| Often | 9 | 2.9 |
| Always | 301 | 97.1 |
| **Amount of salt consumption** |  |  |
| Too much | 17 | 5.5 |
| Just the right amount | 293 | 94.5 |
| **Perception regarding lowering salt in diet** |  |  |
| Very important | 237 | 76.5 |
| Somewhat important | 59 | 19.0 |
| Not at all important | 14 | 4.5 |
| **Vigorous activity during work** |  |  |
| Yes | 42 | 13.5 |
| No | 268 | 86.5 |
| **Moderate activity during work** |  |  |
| Yes | 50 | 16.1 |
| No | 260 | 83.9 |
| **BMI (kg/m<sup>2</sup>)** |  |  |
| Underweight (<18.5) | 14 | 4.5 |
| Normal (18.5 - 24.9) | 271 | 87.4 |
| Overweight (≥25.0) | 23 | 7.4 |
| Obesity (≥30.0) | 2 | 0.6 |
| **Family History of HTN** |  |  |
| Yes | 51 | 16.5 |
| No | 259 | 83.5 |
| **Personal History of HTN** |  |  |
| Yes | 7 | 2.3 |
| No | 303 | 97.7 |
| **Parental history of MI (age <60)** |  |  |
| Yes | 6 | 1.9 |
| No | 304 | 98.1 |
| **Daily Fruits Serving** |  |  |
| 0-1 | 264 | 85.2 |
| 1-3 | 39 | 12.6 |
| 3-5 | 7 | 2.3 |
| **Daily Vegetables Serving** |  |  |
| 0-1 | 215 | 69.4 |
| 1-3 | 69 | 22.3 |
| 3-5 | 26 | 8.4 |
| **Hours spent sitting daily** |  |  |
| 0-3 | 258 | 83.2 |
| 4-6 | 36 | 11.6 |
| 7-8 | 16 | 5.2 |
| **Continuous Variables** | **Mean** | **Std. Deviation** |
| **Systolic Blood Pressure** | 131.11 | 18.616 |
| **Diastolic Blood Pressure** | 78.63 | 13.122 |

**Table 2:**
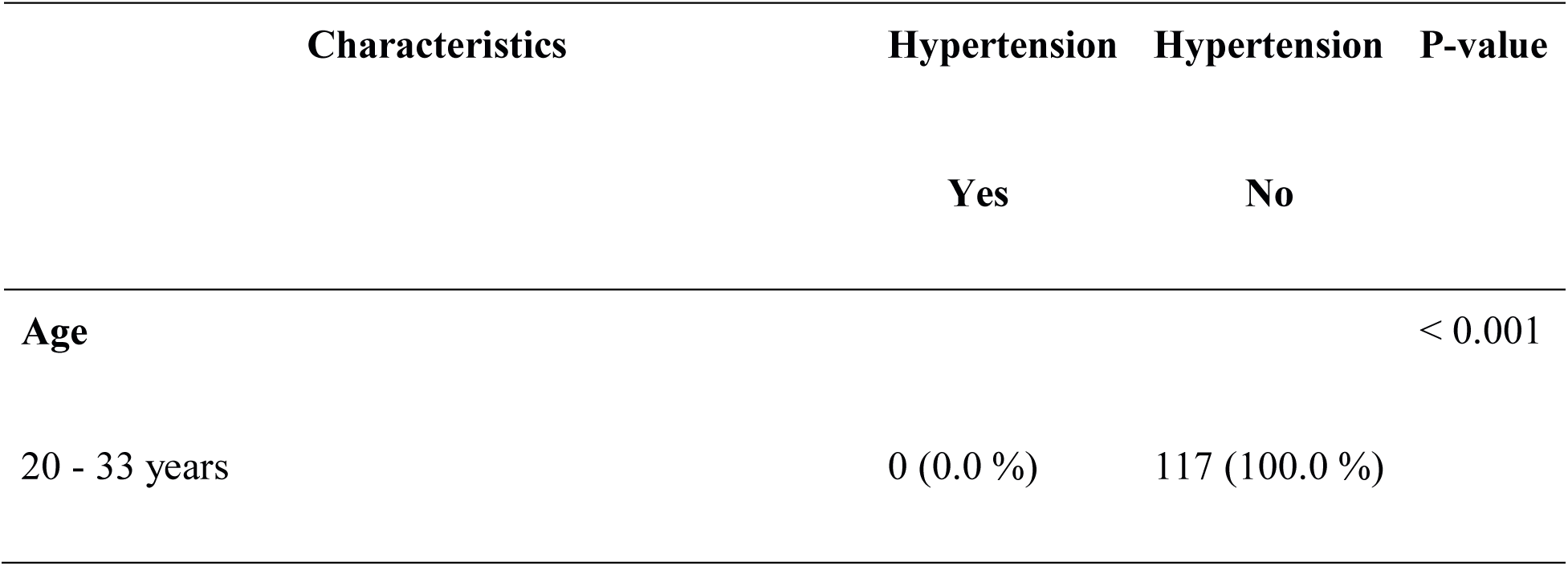
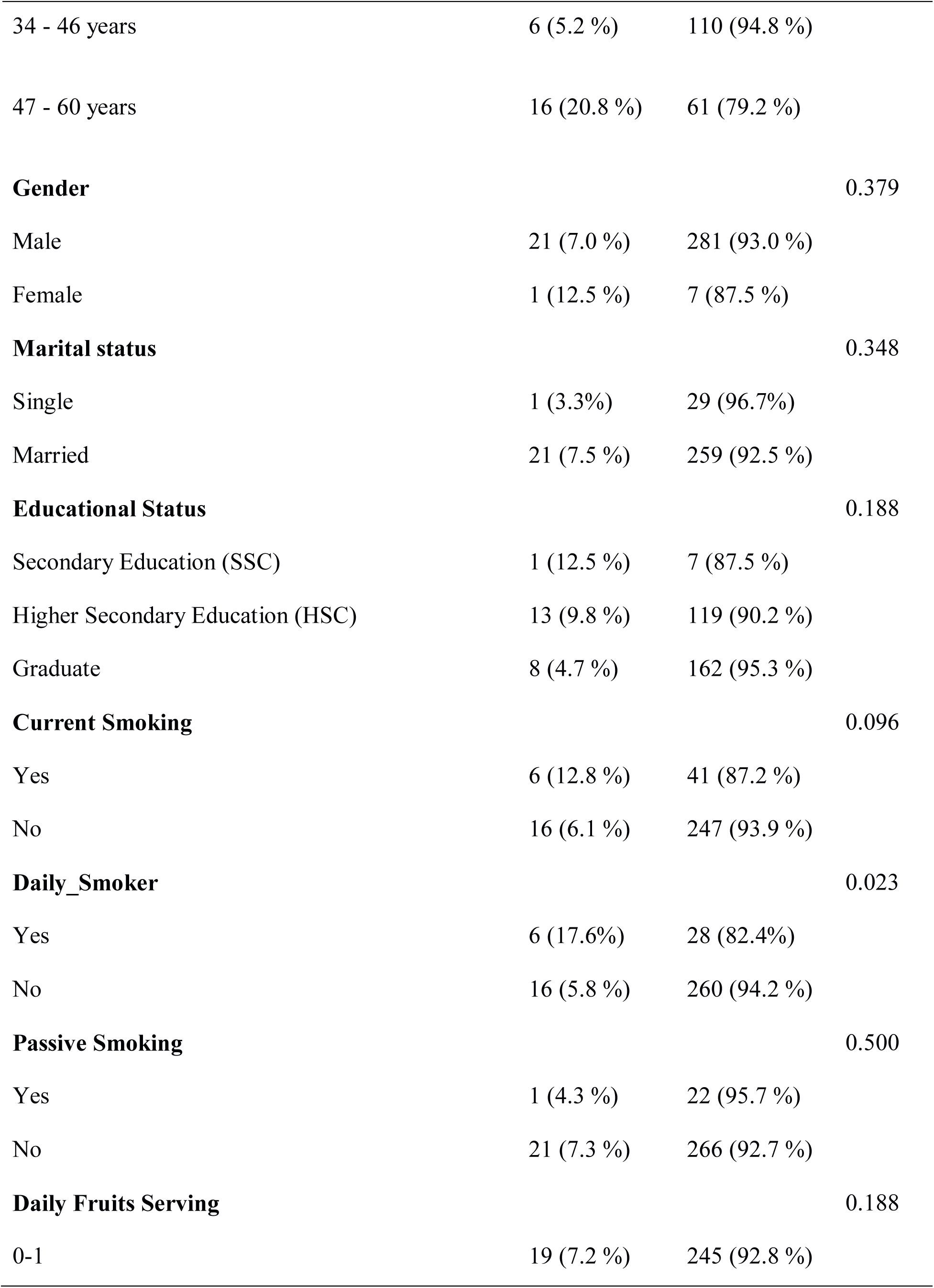
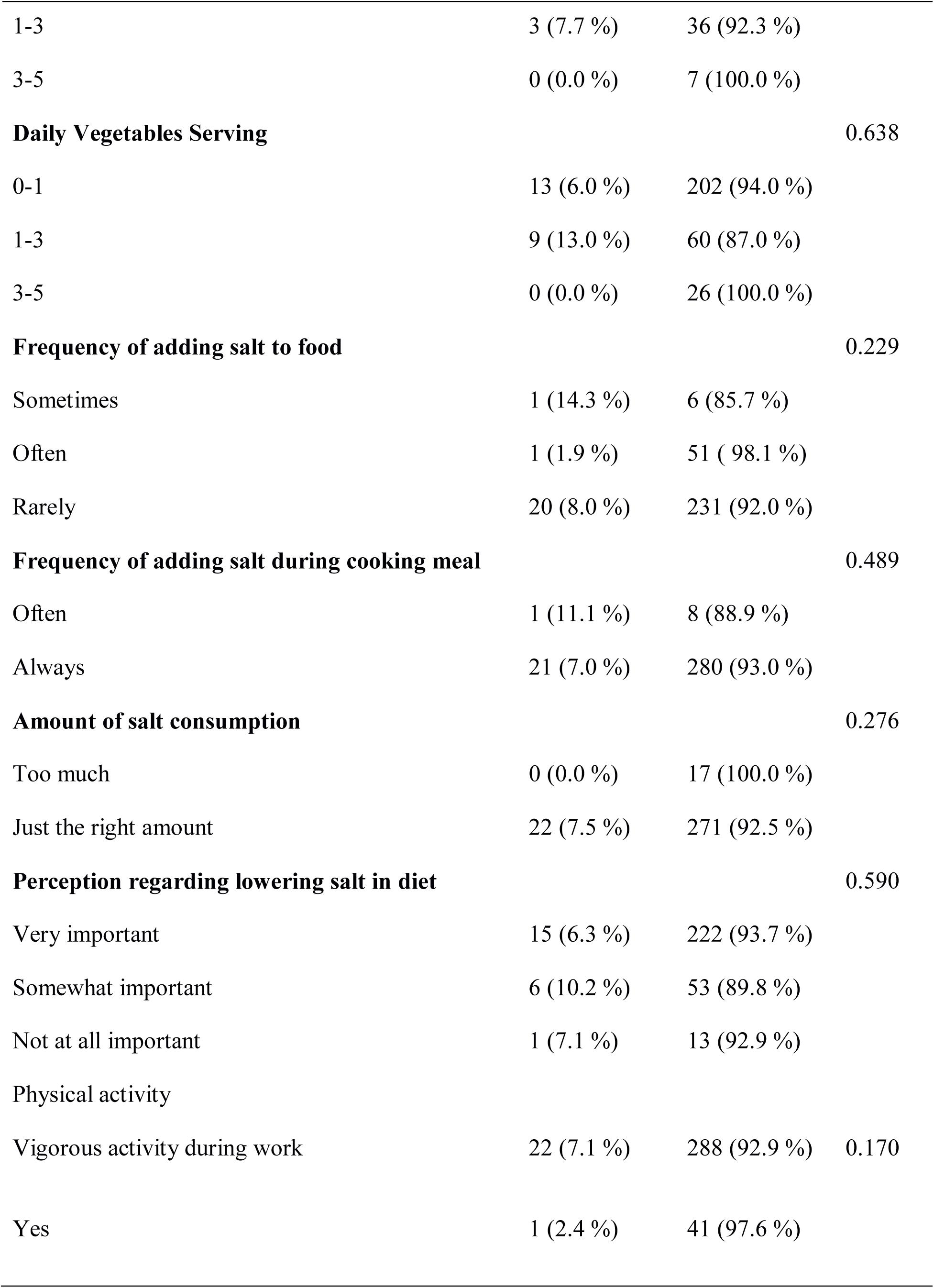
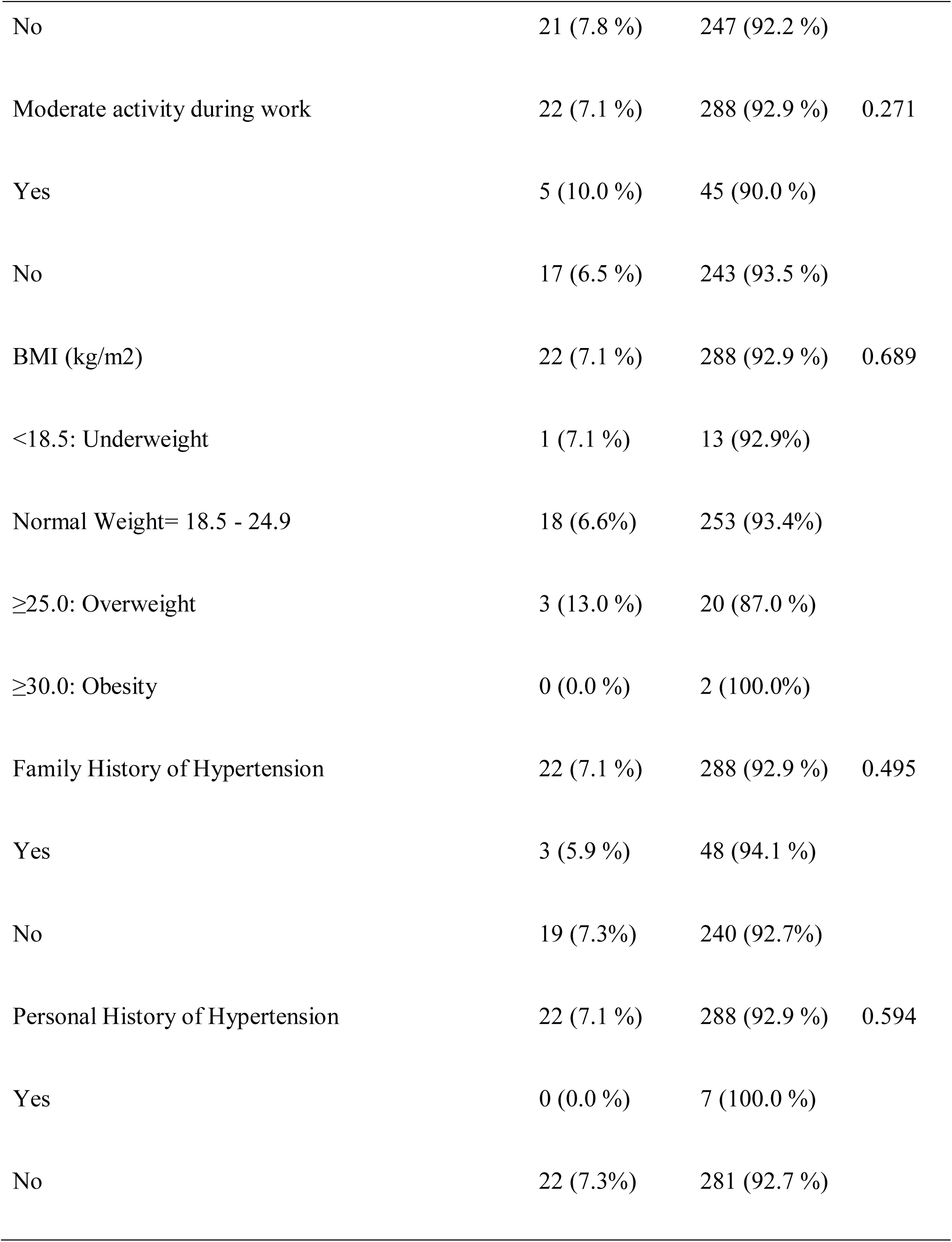
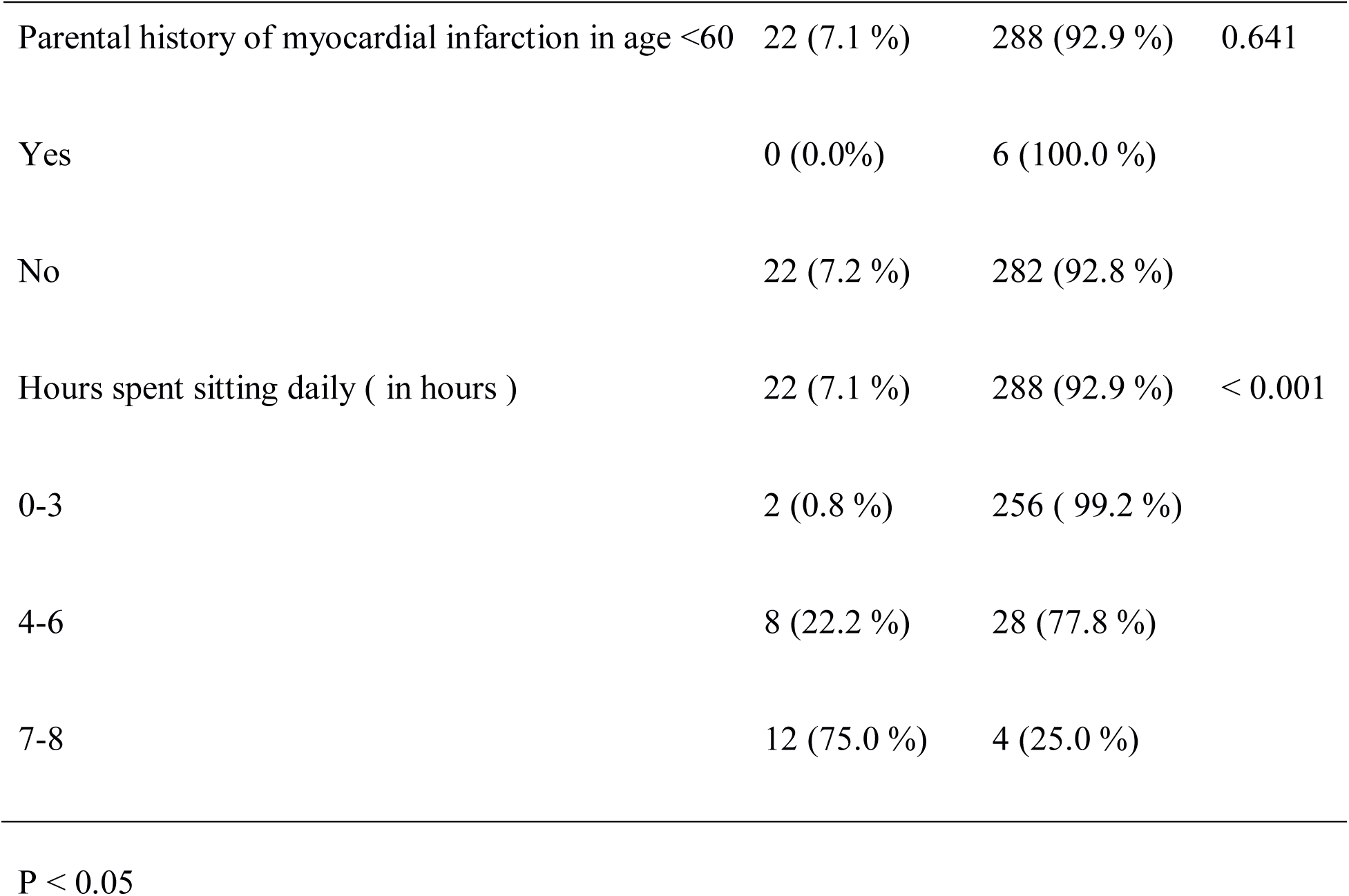
Differences in the baseline characteristics of the study population between the non-hypertension and hypertension groups.

| Characteristics | Hypertension | Hypertension | P-value |
| --- | --- | --- | --- |
|  | Yes | No |  |
| **Age** |  |  | < 0.001 |
| 20 - 33 years | 0 (0.0 %) | 117 (100.0 %) |  |
| 34 - 46 years | 6 (5.2 %) | 110 (94.8 %) |  |
| 47 - 60 years | 16 (20.8 %) | 61 (79.2 %) |  |
| **Gender** |  |  | 0.379 |
| Male | 21 (7.0 %) | 281 (93.0 %) |  |
| Female | 1 (12.5 %) | 7 (87.5 %) |  |
| **Marital status** |  |  | 0.348 |
| Single | 1 (3.3%) | 29 (96.7%) |  |
| Married | 21 (7.5 %) | 259 (92.5 %) |  |
| **Educational Status** |  |  | 0.188 |
| Secondary Education (SSC) | 1 (12.5 %) | 7 (87.5 %) |  |
| Higher Secondary Education (HSC) | 13 (9.8 %) | 119 (90.2 %) |  |
| Graduate | 8 (4.7 %) | 162 (95.3 %) |  |
| **Current Smoking** |  |  | 0.096 |
| Yes | 6 (12.8 %) | 41 (87.2 %) |  |
| No | 16 (6.1 %) | 247 (93.9 %) |  |
| **Daily_Smoker** |  |  | 0.023 |
| Yes | 6 (17.6%) | 28 (82.4%) |  |
| No | 16 (5.8 %) | 260 (94.2 %) |  |
| **Passive Smoking** |  |  | 0.500 |
| Yes | 1 (4.3 %) | 22 (95.7 %) |  |
| No | 21 (7.3 %) | 266 (92.7 %) |  |
| **Daily Fruits Serving** |  |  | 0.188 |
| 0-1 | 19 (7.2 %) | 245 (92.8 %) |  |
| 1-3 | 3 (7.7 %) | 36 (92.3 %) |  |
| 3-5 | 0 (0.0 %) | 7 (100.0 %) |  |
| **Daily Vegetables Serving** |  |  | 0.638 |
| 0-1 | 13 (6.0 %) | 202 (94.0 %) |  |
| 1-3 | 9 (13.0 %) | 60 (87.0 %) |  |
| 3-5 | 0 (0.0 %) | 26 (100.0 %) |  |
| **Frequency of adding salt to food** |  |  | 0.229 |
| Sometimes | 1 (14.3 %) | 6 (85.7 %) |  |
| Often | 1 (1.9 %) | 51 ( 98.1 %) |  |
| Rarely | 20 (8.0 %) | 231 (92.0 %) |  |
| **Frequency of adding salt during cooking meal** |  |  | 0.489 |
| Often | 1 (11.1 %) | 8 (88.9 %) |  |
| Always | 21 (7.0 %) | 280 (93.0 %) |  |
| **Amount of salt consumption** |  |  | 0.276 |
| Too much | 0 (0.0 %) | 17 (100.0 %) |  |
| Just the right amount | 22 (7.5 %) | 271 (92.5 %) |  |
| **Perception regarding lowering salt in diet** |  |  | 0.590 |
| Very important | 15 (6.3 %) | 222 (93.7 %) |  |
| Somewhat important | 6 (10.2 %) | 53 (89.8 %) |  |
| Not at all important | 1 (7.1 %) | 13 (92.9 %) |  |
| **Physical activity** |  |  |  |
| Vigorous activity during work | 22 (7.1 %) | 288 (92.9 %) | 0.170 |
| Yes | 1 (2.4 %) | 41 (97.6 %) |  |
| No | 21 (7.8 %) | 247 (92.2 %) |  |
| Moderate activity during work | 22 (7.1 %) | 288 (92.9 %) | 0.271 |
| Yes | 5 (10.0 %) | 45 (90.0 %) |  |
| No | 17 (6.5 %) | 243 (93.5 %) |  |
| BMI (kg/m2) | 22 (7.1 %) | 288 (92.9 %) | 0.689 |
| <18.5: Underweight | 1 (7.1 %) | 13 (92.9%) |  |
| Normal Weight= 18.5 - 24.9 | 18 (6.6%) | 253 (93.4%) |  |
| ≥25.0: Overweight | 3 (13.0 %) | 20 (87.0 %) |  |
| ≥30.0: Obesity | 0 (0.0 %) | 2 (100.0%) |  |
| Family History of Hypertension | 22 (7.1 %) | 288 (92.9 %) | 0.495 |
| Yes | 3 (5.9 %) | 48 (94.1 %) |  |
| No | 19 (7.3%) | 240 (92.7%) |  |
| Personal History of Hypertension | 22 (7.1 %) | 288 (92.9 %) | 0.594 |
| Yes | 0 (0.0 %) | 7 (100.0 %) |  |
| No | 22 (7.3%) | 281 (92.7 %) |  |
| Parental history of myocardial infarction in age <60 | 22 (7.1 %) | 288 (92.9 %) | 0.641 |
| Yes | 0 (0.0%) | 6 (100.0 %) |  |
| No | 22 (7.2 %) | 282 (92.8 %) |  |
| Hours spent sitting daily ( in hours ) | 22 (7.1 %) | 288 (92.9 %) | < 0.001 |
| 0-3 | 2 (0.8 %) | 256 ( 99.2 %) |  |
| 4-6 | 8 (22.2 %) | 28 (77.8 %) |  |
| 7-8 | 12 (75.0 %) | 4 (25.0 %) |  |

P < 0.05

**Table 3:**
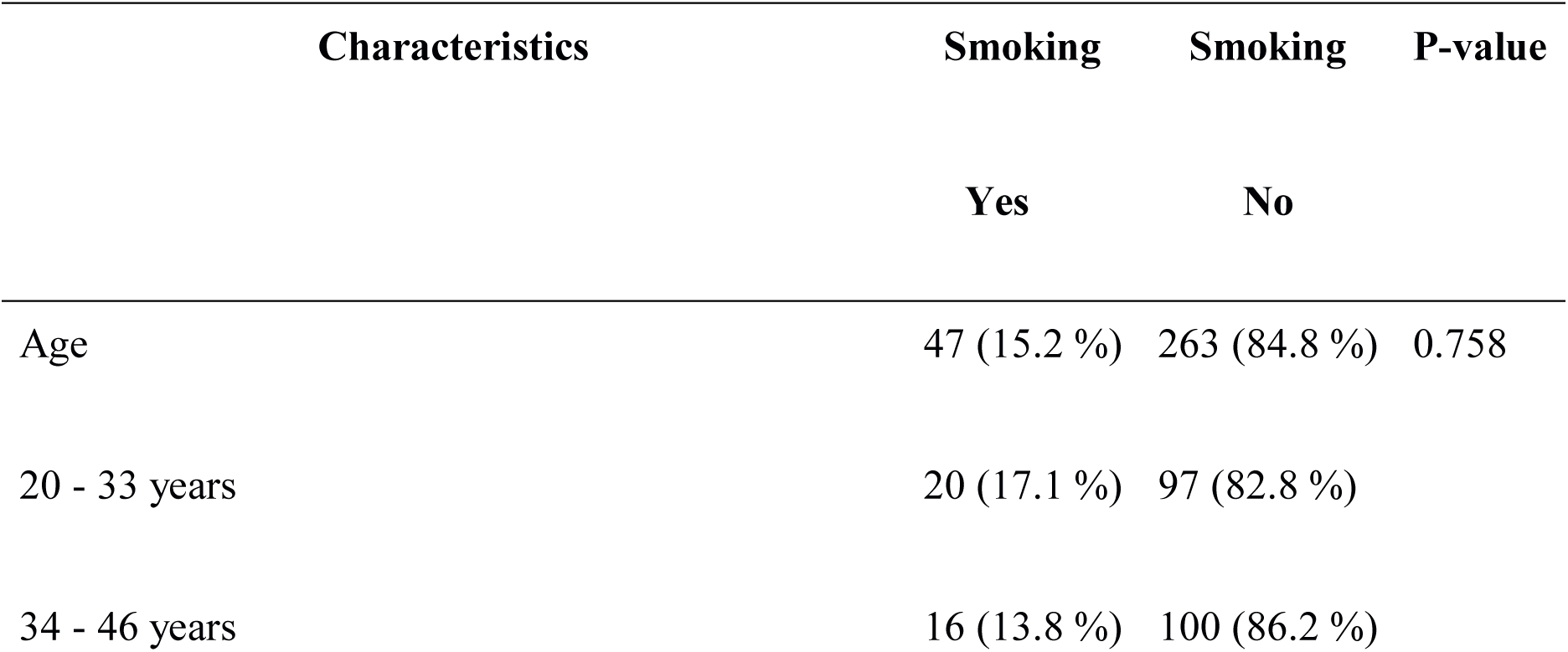
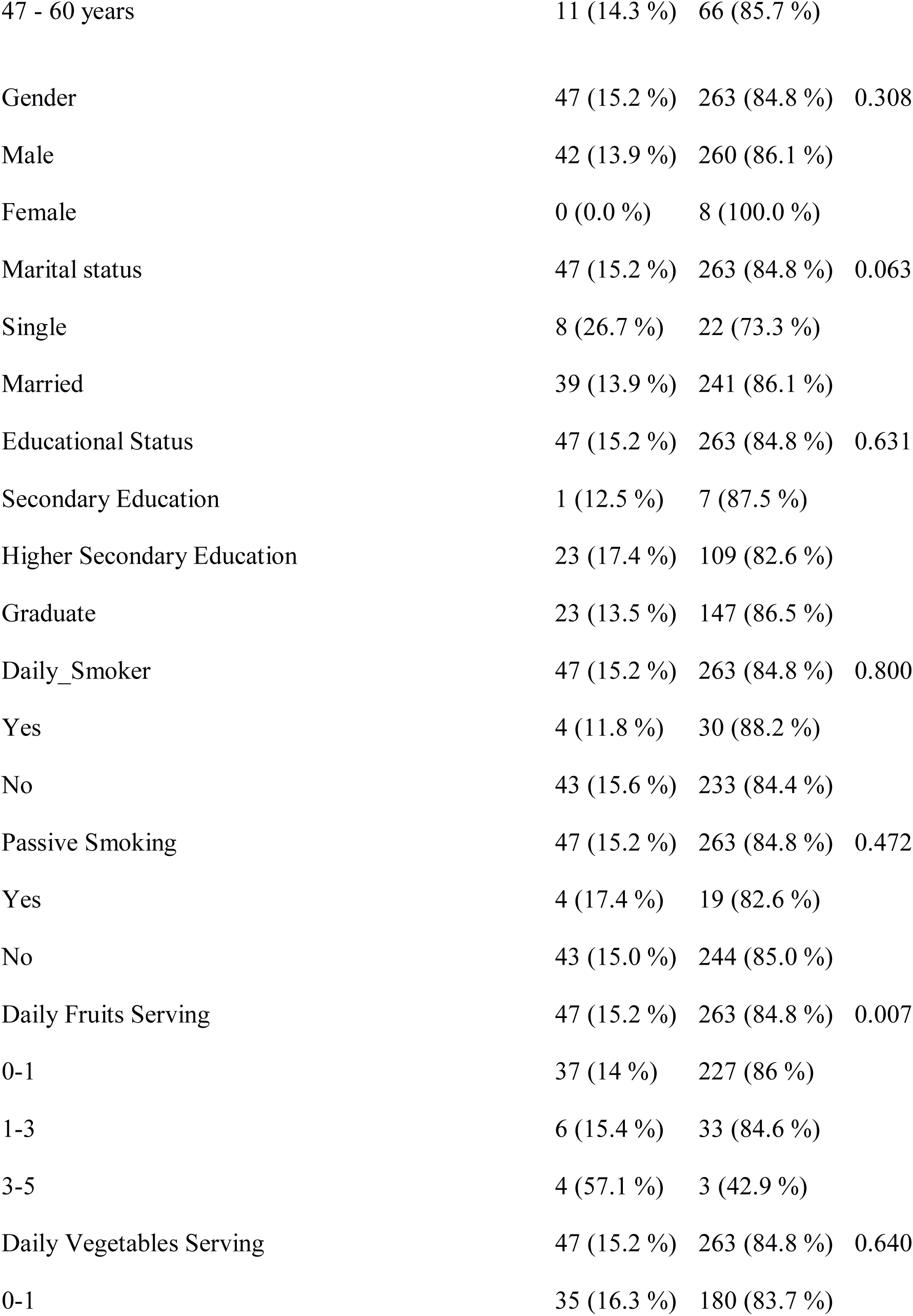
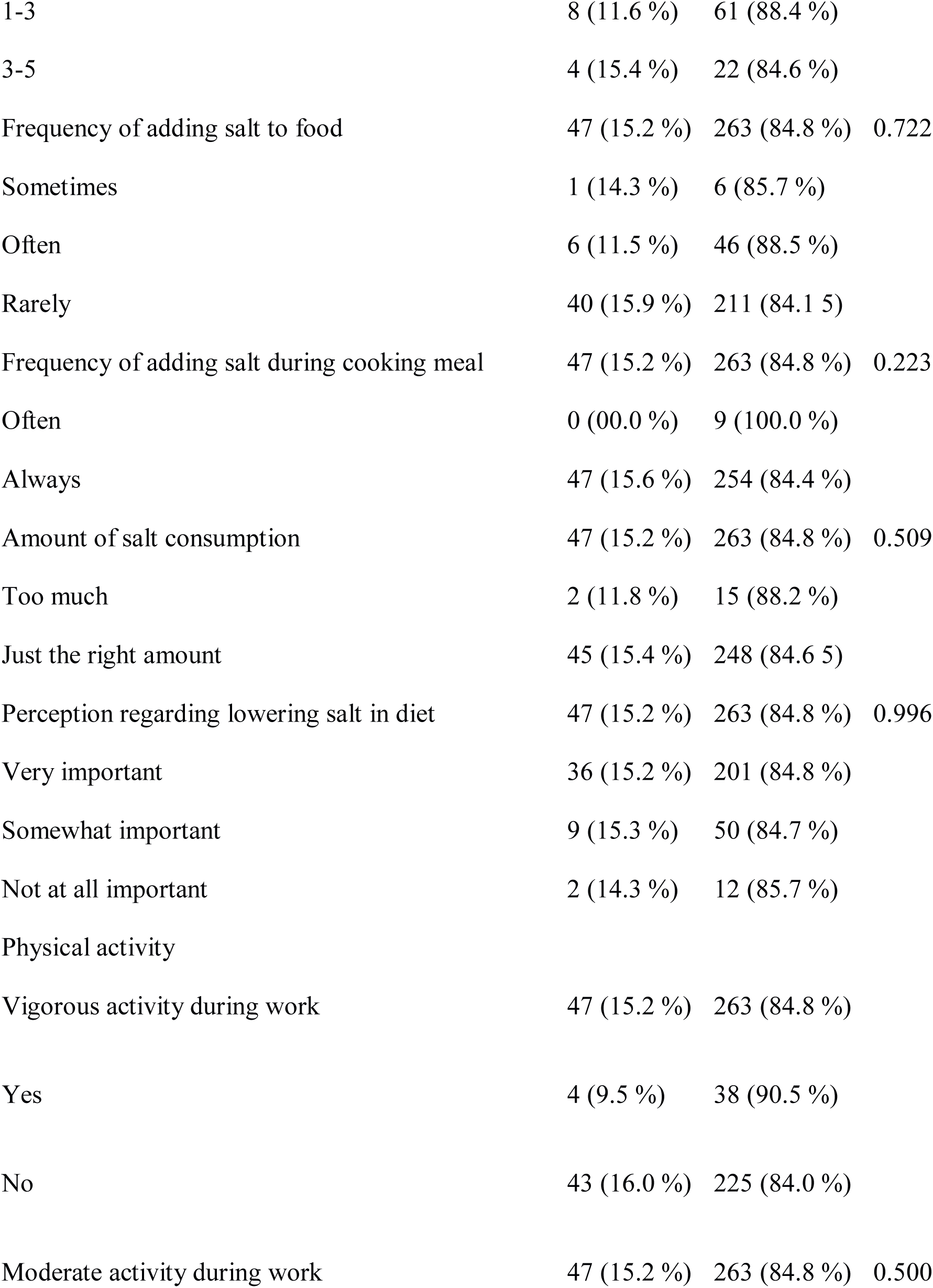
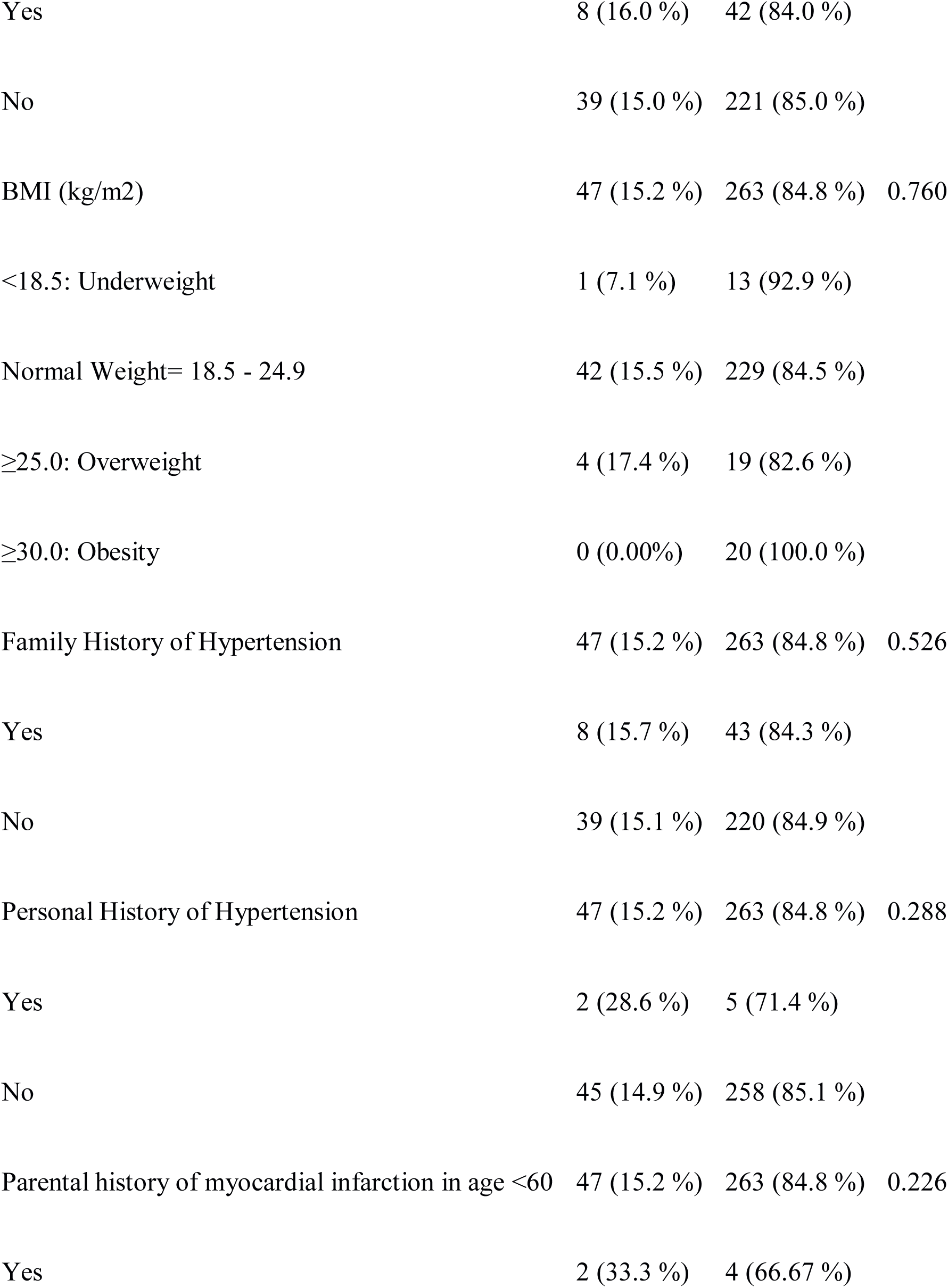
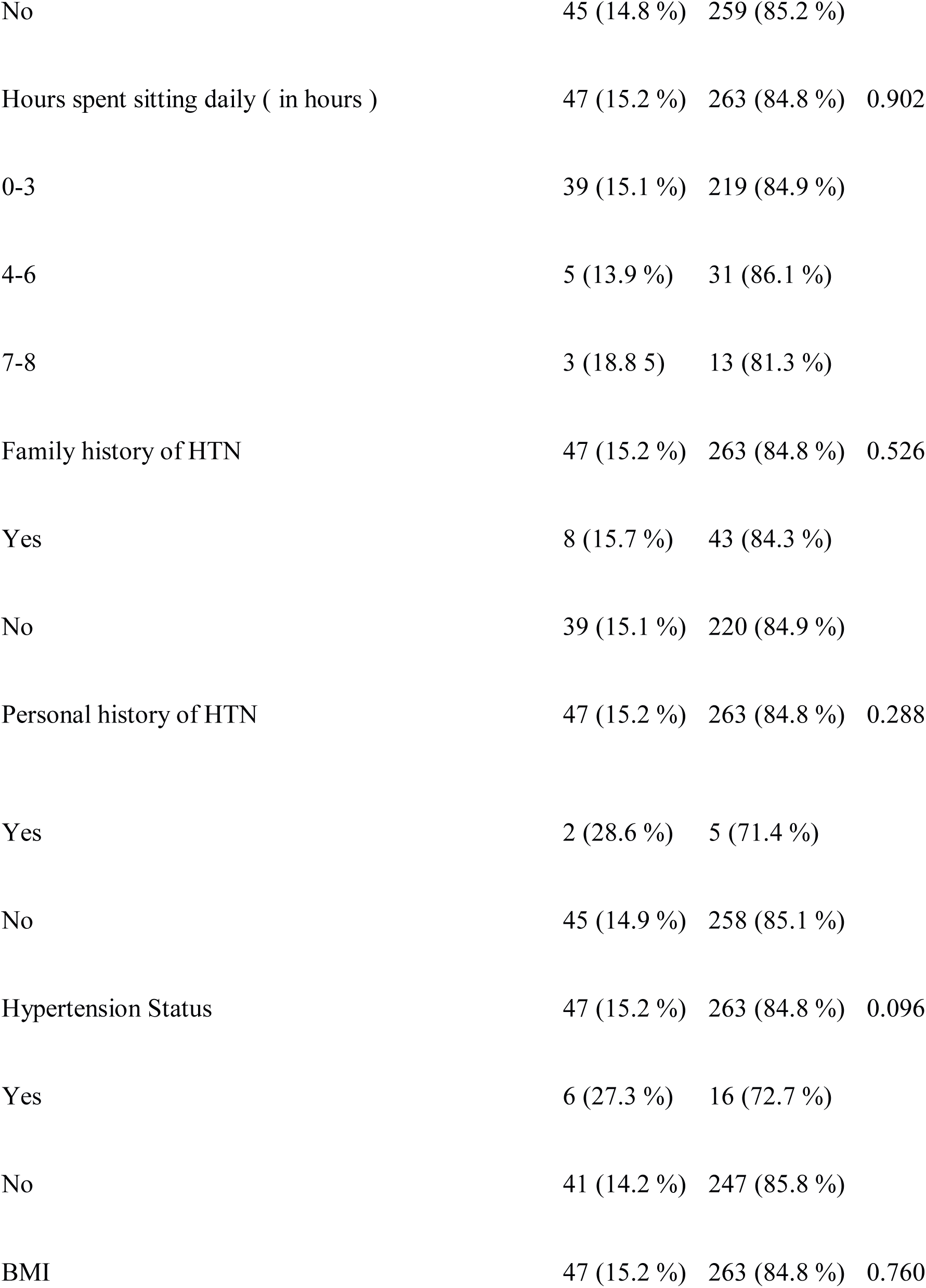
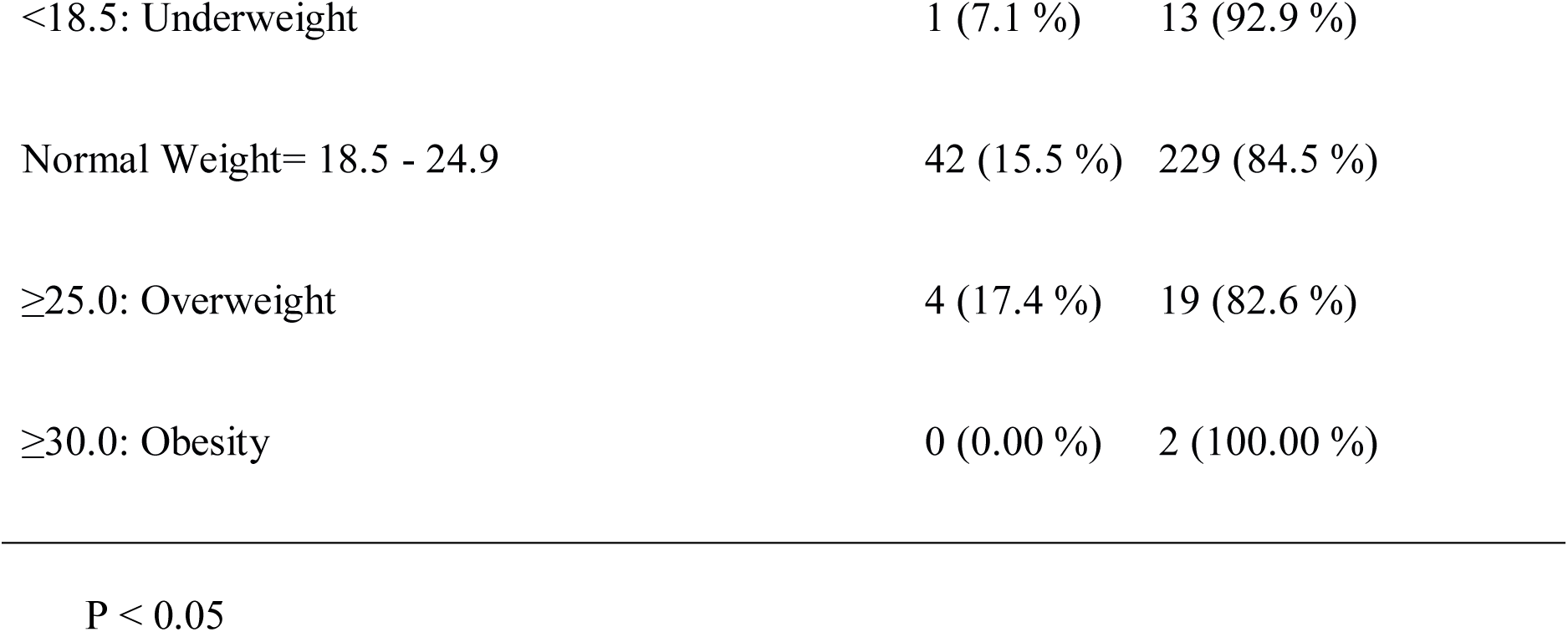
Differences in the baseline characteristics of the study population between the non-smoker and smoker groups.

| Characteristics | Smoking | Smoking | P-value |
| --- | --- | --- | --- |
|  | Yes | No |  |
| Age | 47 (15.2 %) | 263 (84.8 %) | 0.758 |
| 20 - 33 years | 20 (17.1 %) | 97 (82.8 %) |  |
| 34 - 46 years | 16 (13.8 %) | 100 (86.2 %) |  |
| 47 - 60 years | 11 (14.3 %) | 66 (85.7 %) |  |
| Gender | 47 (15.2 %) | 263 (84.8 %) | 0.308 |
| Male | 42 (13.9 %) | 260 (86.1 %) |  |
| Female | 0 (0.0 %) | 8 (100.0 %) |  |
| Marital status | 47 (15.2 %) | 263 (84.8 %) | 0.063 |
| Single | 8 (26.7 %) | 22 (73.3 %) |  |
| Married | 39 (13.9 %) | 241 (86.1 %) |  |
| Educational Status | 47 (15.2 %) | 263 (84.8 %) | 0.631 |
| Secondary Education | 1 (12.5 %) | 7 (87.5 %) |  |
| Higher Secondary Education | 23 (17.4 %) | 109 (82.6 %) |  |
| Graduate | 23 (13.5 %) | 147 (86.5 %) |  |
| Daily_Smoker | 47 (15.2 %) | 263 (84.8 %) | 0.800 |
| Yes | 4 (11.8 %) | 30 (88.2 %) |  |
| No | 43 (15.6 %) | 233 (84.4 %) |  |
| Passive Smoking | 47 (15.2 %) | 263 (84.8 %) | 0.472 |
| Yes | 4 (17.4 %) | 19 (82.6 %) |  |
| No | 43 (15.0 %) | 244 (85.0 %) |  |
| Daily Fruits Serving | 47 (15.2 %) | 263 (84.8 %) | 0.007 |
| 0-1 | 37 (14 %) | 227 (86 %) |  |
| 1-3 | 6 (15.4 %) | 33 (84.6 %) |  |
| 3-5 | 4 (57.1 %) | 3 (42.9 %) |  |
| Daily Vegetables Serving | 47 (15.2 %) | 263 (84.8 %) | 0.640 |
| 0-1 | 35 (16.3 %) | 180 (83.7 %) |  |
| 1-3 | 8 (11.6 %) | 61 (88.4 %) |  |
| 3-5 | 4 (15.4 %) | 22 (84.6 %) |  |
| Frequency of adding salt to food | 47 (15.2 %) | 263 (84.8 %) | 0.722 |
| Sometimes | 1 (14.3 %) | 6 (85.7 %) |  |
| Often | 6 (11.5 %) | 46 (88.5 %) |  |
| Rarely | 40 (15.9 %) | 211 (84.1 %) |  |
| Frequency of adding salt during cooking meal | 47 (15.2 %) | 263 (84.8 %) | 0.223 |
| Often | 0 (00.0 %) | 9 (100.0 %) |  |
| Always | 47 (15.6 %) | 254 (84.4 %) |  |
| Amount of salt consumption | 47 (15.2 %) | 263 (84.8 %) | 0.509 |
| Too much | 2 (11.8 %) | 15 (88.2 %) |  |
| Just the right amount | 45 (15.4 %) | 248 (84.6 %) |  |
| Perception regarding lowering salt in diet | 47 (15.2 %) | 263 (84.8 %) | 0.996 |
| Very important | 36 (15.2 %) | 201 (84.8 %) |  |
| Somewhat important | 9 (15.3 %) | 50 (84.7 %) |  |
| Not at all important | 2 (14.3 %) | 12 (85.7 %) |  |
| Physical activity |  |  |  |
| Vigorous activity during work | 47 (15.2 %) | 263 (84.8 %) |  |
| Yes | 4 (9.5 %) | 38 (90.5 %) |  |
| No | 43 (16.0 %) | 225 (84.0 %) |  |
| Moderate activity during work | 47 (15.2 %) | 263 (84.8 %) | 0.500 |
| Yes | 8 (16.0 %) | 42 (84.0 %) |  |
| No | 39 (15.0 %) | 221 (85.0 %) |  |
| BMI (kg/m2) | 47 (15.2 %) | 263 (84.8 %) | 0.760 |
| <18.5: Underweight | 1 (7.1 %) | 13 (92.9 %) |  |
| Normal Weight= 18.5 - 24.9 | 42 (15.5 %) | 229 (84.5 %) |  |
| ≥25.0: Overweight | 4 (17.4 %) | 19 (82.6 %) |  |
| ≥30.0: Obesity | 0 (0.00%) | 20 (100.0 %) |  |
| Family History of Hypertension | 47 (15.2 %) | 263 (84.8 %) | 0.526 |
| Yes | 8 (15.7 %) | 43 (84.3 %) |  |
| No | 39 (15.1 %) | 220 (84.9 %) |  |
| Personal History of Hypertension | 47 (15.2 %) | 263 (84.8 %) | 0.288 |
| Yes | 2 (28.6 %) | 5 (71.4 %) |  |
| No | 45 (14.9 %) | 258 (85.1 %) |  |
| Parental history of myocardial infarction in age <60 | 47 (15.2 %) | 263 (84.8 %) | 0.226 |
| Yes | 2 (33.3 %) | 4 (66.67 %) |  |
| No | 45 (14.8 %) | 259 (85.2 %) |  |
| Hours spent sitting daily ( in hours ) | 47 (15.2 %) | 263 (84.8 %) | 0.902 |
| 0-3 | 39 (15.1 %) | 219 (84.9 %) |  |
| 4-6 | 5 (13.9 %) | 31 (86.1 %) |  |
| 7-8 | 3 (18.8 %) | 13 (81.3 %) |  |
| Family history of HTN | 47 (15.2 %) | 263 (84.8 %) | 0.526 |
| Yes | 8 (15.7 %) | 43 (84.3 %) |  |
| No | 39 (15.1 %) | 220 (84.9 %) |  |
| Personal history of HTN | 47 (15.2 %) | 263 (84.8 %) | 0.288 |
| Yes | 2 (28.6 %) | 5 (71.4 %) |  |
| No | 45 (14.9 %) | 258 (85.1 %) |  |
| Hypertension Status | 47 (15.2 %) | 263 (84.8 %) | 0.096 |
| Yes | 6 (27.3 %) | 16 (72.7 %) |  |
| No | 41 (14.2 %) | 247 (85.8 %) |  |
| BMI | 47 (15.2 %) | 263 (84.8 %) | 0.760 |

|  |  |  |
| --- | --- | --- |
| <18.5: Underweight | 1 (7.1 %) | 13 (92.9 %) |
| Normal Weight= 18.5 - 24.9 | 42 (15.5 %) | 229 (84.5 %) |
| ≥25.0: Overweight | 4 (17.4 %) | 19 (82.6 %) |
| ≥30.0: Obesity | 0 (0.00 %) | 2 (100.00 %) |

P < 0.05

### Results and Findings

Table 1 presents the descriptive statistics of the population, where the distributions of the categorical and continuous variables are considered. Consequently, it ensures that 37.7% of the sample pertains to the age brackets between 20-33 years and 37.4% pertain to the 34-46-year age bracket, while the rest of the subjects are distributed within the 47-60-year age bracket. The sample is also highly male-dominated, with 97.4% males, against a meager 2.6% females. The majority of the respondents were married, 90.3%. A greater number of the participants fall into higher secondary education with 54.8% being graduates, 42.6% with higher secondary, and 2.6% with secondary education. Currently, 15.2% smoke, and 11% are smoking daily. Only 7.4% reported passive smoking. For dietary behavior, 81% of the participants reported that they “rarely” add salt to food, whereas 97.1% “always” add salt while cooking, and 94.5% claim that their salt consumption is “just the right amount.” Moreover, 76.5% indicated that it is “very important” to reduce salt in their diets. For physical activities, 13.5% of the participants performed vigorous activities at work and 16.1% performed moderate activities. The majority of participants have a normal BMI, 87.4%, while a small proportion of the subjects reported being underweight 4.5%, or overweight 7.4%. Family history of hypertension is given by 16.5%, and only 2.3% of the subjects have been diagnosed with hypertension. Family history of myocardial infarction before age 60 is seldom present, 1.9%. The majority of participants consume 0-1 daily serving of fruits, 85.2%, and vegetables, 69.4%. The sedentary behavior: 83.2% of the respondents spend 0-3 hours sitting per day. Continuous variables are represented by the mean systolic blood pressure of 131.11 mmHg ± 18.616 and a mean diastolic blood pressure of 78.63 mmHg ± 13.122.

Table 2 compares the baseline characteristics of those participants with and those without hypertension, demonstrating statistically significant differences between the groups regarding age, daily smoking, and hours of sitting daily. It shows a statistically significant difference in age distribution, p < 0.001; the prevalence of hypertension was highly marked in the group of participants falling within an age group of 47-60 years (20.8%) when compared to the younger groups. It was also shown that the proportion of daily smokers with hypertension was considerably higher −17.6 % as opposed to 5.8% among nondaily smokers at a p-value of 0.023. The number of hours sitting each day showed a strong association: p < 0.001, indicating that individuals who sat for 7 to 8 hours had hypertension at a rate of 75.0%, while those who sat for 0-3 hours recorded a hypertension prevalence of only 0.8%. Whereas there were no significant differences for gender, marital status, educational level, and most of the items on lifestyle factors, a higher percentage of overweight people have hypertension, 13.0%, compared to those within the normal BMI, 6.6%. Family history of hypertension, personal history of myocardial infarction, salt intake, and physical activity are also not significantly associated with hypertension. These findings further establish the contribution of age, smoking, and sedentary activity to hypertension in this study population.

Table 3 compares the baseline characteristics between smokers and non-smokers. Distribution of smokers according to age was: 20-33 years, 17.1%; 34-46 years, 13.8%; 47-60 years, 14.3%, without statistical difference for smokers and non-smokers: p = 0.758. There were no gender differences, p = 0.308, as 13.9% of males were recorded to smoke, while no female smokers were noted. Marital status was marginal to significance, p = 0.063, while 26.7% of single participants smoke versus 13.9% of married ones. However, educational status was insignificant, p = 0.631; the highest smoking prevalence in those with higher secondary education comprised 17.4%. The daily smokers comprised 11.8% of the smoker group, and 17.4% practiced passive smoking; however, both factors are nonsignificant, p = 0.800 and p = 0.472, respectively. However, a significant difference in the daily serving of fruits, p = 0.007, where smokers take 3- 5 servings more than nonsmokers, accounting for 57.1%, no vegetable salt consumption and physical activity differences were observed. Distribution of BMI indicated that more overweight were among the smokers than nonsmokers by a percentage of 17.4%, but it was not at a significant level of p = 0.760. Also, the family and personal histories of hypertension, and parental myocardial infarction were not different across the groups significantly. The hours per day spent sitting were not associated, having a p-value of 0.902. Hypertension status is not different as well, though it is close to statistical significance with a p-value of 0.096, having 27.3% of smokers having hypertension status as against 14.2% of nonsmokers. In a nutshell, all the demographic, lifestyle, and health-related variables have been represented in the table, with significant differences present only in fruit consumption.

## Discussion

It was a comprehensive study on the prevalence and major drivers of hypertension among employees of a beverage company based in Dhaka. The present study, therefore, encompasses all management and non-management personnel within the age bracket of 20-60 years. A cross-sectional design was applied, and the measurement of blood pressure was done based on a uniform standard procedure. A detailed survey was conducted by applying the World Health Organization’s STEPS approach to the surveillance of noncommunicable diseases. The survey instrument, after refinement through a pilot study, comprises items on socio-demographic information and information on cardiovascular risk factors. Composition of Participants The participants in this study constituted 97.4% males and 2.6% females. The participants fell roughly equally within the age brackets of 20-33, 34-46, and 47-60 years, which also represents a good adult working population. Most of the respondents were married, at 90.3%, while 84.8% did not have a smoking habit, even though 15.2% were smokers and 7.4% passive smokers. History of hypertension was very low and diagnosed only in 7.1%, while 97.7% reported no history of hypertension.

This was reflected in the practices of salt consumption: 97.1% of the respondents added salt during cooking, and 94.5% mentioned that they consumed the “right” amount of salt. However, a high proportion of 76.5% perceived the reduction of salt intake as important. These results are expected from the national dietary habits in Bangladesh, where adding salt during the preparation of food is a very common practice. Despite all this, the prevalence of hypertension again was found to be rather low, perhaps reflecting a general characteristic of health among this relatively younger, working population.

Physical activity was low, with only 13.5% reporting vigorous activity at work and 16.1% having moderate activity. Furthermore, 83.2% reported sitting for 0-3 hours daily, which would show a fairly active lifestyle, though this might well be negated by the lack of strenuous physical activities. They still kept a good distribution in terms of BMI, with 87.4% falling within the normal range and only 7.4% overweight.

There were significant differences in age and hours sitting daily between the two groups: hypertensive and non-hypertensive. Hypertension was most prevalent among participants who were of older ages, in the ranges between 47-60 years; this demographic also had higher rates of prevalence among those who spent more time sitting daily. These findings agree with well-recognized risk factors for hypertension, such as age and physical inactivity. Except for age, no significant associations were found concerning gender, marital status, and smoking habits; although, daily smokers were somewhat more likely to be hypertensive.

During this comparison of smokers versus non-smokers, no significance is realized in most demographic variables, including age, gender, marital status, and level of education. In any case, the intake of fruits was significantly lower among smokers because a greater number of the respondents reported intakes of not up to 1-3 servings each day. It also falls in line with past literature where smoking is related to poor dietary propensities and accordingly poor well-being of smokers.

Overall, findings suggest that though the general population engages in healthy behavior, the population subgroups are still at risk of disease, like hypertension, which would serve to disadvantage subgroups such as the elderly and smokers. This underlines the necessity for targeted lifestyle modification interventions on increasing physical activity and improving dietary habits to reduce the risk of diseases. Also, data indicate the need for public health campaigns on smoking reduction and healthy eating patterns in the workplace, where sitting is more common.

These reviewed studies with the literature referred to in them suggest a certain consistency of the observed patterns related to the smoking factor in the aspect of its relationship with hypertension, notwithstanding that in these studies, both the population and methodologies applied differ.

Some review says that Li et al., 2017; Wang et al., 2020 explained a higher prevalence of hypertension among former smokers compared to current smokers. Li et al. (2017) and Wang et al. (2020) have reported higher blood pressure in former smokers. In this regard, John et al. (2006) have shown increased risks of moderate to severe hypertension among former smokers, much more pronounced among obese ones. Similarly, ex-smokers have been found to show higher odds of hypertension across studies.

Li et al. also pointed out that current smokers had lower SBP as compared to ex-smokers, and that agrees with the findings presented by Wang et al. (2020), whereby regular smokers showed lower SBP and DBP. That is the exact trend which was observed in this analysis the current smokers had lower blood pressure levels as compared to the nonsmokers and ex-smokers. Such facts are further supported by Li et al. (2017), who mentions that smoking in pack-years was associated with dose-dependently increasing SBP, DBP, and mean arterial pressure in agreement with other studies that suggest a higher risk of hypertension associated with longer exposure to smoking.

The other most common factor that has been largely present in the majority of findings is the interaction between smoking, BMI, and hypertension. Yao et al. (2020) demonstrated that current smoking was a strong predictor of hypertension among persons with normal weight status, but BMI remained a major predictor across smoking status. On the other hand, John et al. (2006) also presented the same interaction in the review findings, where such findings are parallel to the present study, where both smoking and BMI modify the risk of hypertension.

Wu et al., 2017, however, did show gender differences, especially passive smoke effects on blood pressure, where a higher prevalence of hypertension in women exposed to passive smoke. The above, therefore, shows wider gender-based differences in smoking and hypertension, as reflected by Mehboudi et al., 2017, and other referenced studies that stratify the prevalence or risk of hypertension according to gender.

The interaction between smoking and alcohol consumption was important, further confirmed by Wang et al. (2020); indeed, the interaction of alcohol with smoking was associated with increased blood pressure in men. These crossing findings explain how smoking, apart from the rest of the elements that concern gender, BMI, and alcohol consumption, is entwined in the regulation of blood pressure and the rise of risk for hypertension.

Regarding gender distribution, this primary study, and that done by Kim et al., in 2018 was predominantly male, because 97.4% of the subjects in the primary study were males. In this regard, Kim et al. have also stated a higher incidence of hypertension for men than for women, 8.6% versus 2.8%, respectively, explaining the gender-related smoking trends.

Both studies showed the association between smoking and hypertension to be statistically significant. While the main study presented only a marginal increase in the hypertensive state of smokers, Kim et al., 2018 did observe that continued smokers reported a higher prevalence of hypertension compared to never-smokers: 8.3% versus 4.7%, respectively. Other lifestyle factors covered were physical activity and dietary habits. In the main study, 83.2% were noted to have low levels of physical activity, sitting for 0-3 hours daily. Lifestyle factors such as BMI and alcohol consumption were identified in the analysis as some of the more important variables in hypertension risk, which is supported by research work where smokers tended to show poorer dietary habits, with lower fruit consumption Okubo et al. (2002).

Both studies identify public health interventions for both the elderly and smoking populations. The findings confirm that one of the major priorities is a reduction in the risk factors of hypertension by smoking and its related lifestyle factors; thus, lifestyle changes among current smokers would greatly reduce the prevalence of hypertension, as suggested by Kim et al. (2018) and Okubo et al. (2002). Overall, the findings of the present study support the findings of the previous studies in establishing an intricate relationship between smoking, hypertension, and several modifying factors.

Overall, the findings indicate that the overall healthy behavior of the population may mask some subgroups, especially older people and smokers, who could be more vulnerable to conditions like hypertension. This therefore calls for targeted lifestyle modification interventions in terms of increasing physical activity and improving dietary habits to minimize the risks of such conditions. These data further highlight the need for public health campaigns on smoking reduction and healthier eating patterns, especially in the workplace where sedentary behaviors prevail.

These studies, like the literature that was referenced, shared some trends that linked smoking and hypertension together, but with different population groups and varying methodologies.

Several studies have found higher proportions of former smokers being hypertensive compared to current smokers, such as Li et al. (2017) and Wang et al. (2020). This agrees with the results of this analysis. Li et al. (2017) and Wang et al. (2020) also indicated that former smokers had higher blood pressure, while John et al. (2006) showed further increased risks of moderate to severe hypertension among ex-smokers, with the high risk again predominant among obese persons. That pattern is consistent in studies where ex-smokers tend to have higher odds of hypertension.

Li et al., 2017, indicated that current smokers exhibited lower SBP as compared to ex-smokers, and this was agreed upon by Wang et al. 2020, who noted that regular smokers exhibited lower SBP and DBP. This therefore agrees with the findings of this analysis, in which it was established that current smokers had lower blood pressure compared to non-smokers and ex-smokers. The dose-response relationship reported by Li et al., (2017), where pack-years of smoking were associated with increased SBP, DBP, and MAP, agrees with the previous work that established the duration of smoking as carrying a higher risk for hypertension (Niskanen et al., 2004).

Another factor that is also common is interactions between smoking, body mass index, and hypertension. Yao et al. (2020) reported that current smoking was a significant predictor of hypertension among those with normal weight, while BMI was the most important predictor across all smoking statuses. Similarly, in John et al. (2006), it was mentioned that such interaction exists; findings that run parallel to this present study, both smoking and body mass index modify the risk of hypertension.

Wu et al. (2017) have pointed out gender disparities, especially on the impact of passive smoking on blood pressure, showing a higher prevalence in the case of women being exposed to passive smoke. Indeed, this can be a reflection of larger gender-based disparities related to smoking and hypertension, as reflected by Mehboudi et al. (2017) and other mentioned studies that further stratify the prevalence or risk of hypertension by gender.

Additionally, the synergistic relationship between alcohol intake and smoking as reported by Wang et al. (2020) carries more significant weight for findings in the current analysis where alcohol in concert with smoking was associated with higher levels of blood pressure among men. Overlapping results would then mean that smoking, as well as other factors like BMI, alcohol consumption, and gender, takes part in multiple variables that regulate blood pressure and thereby influence the risk of developing hypertension. This was addressed by Li et al., Wu et al., and Wang et al.

Both the primary study and that of Kim et al. (2018) recorded a male preponderance in their sample population, with 97.4% of the respondents in the primary study being male. Similarly, Kim et al. noted that hypertension was higher among males than females, which agrees with the cigarette smoking trend by gender, at 8.6% versus 2.8%, respectively.

Smoking and hypertension were also observed to share a significant relationship in both studies. Smokers were more likely to have hypertension. For Kim et al., 2018, continued smokers presented with higher rates of hypertension compared to never-smokers, 8.3 percent versus 4.7 percent, respectively. The lifestyle factors included physical activity and dietary habits. In the primary study, 83.2 percent sat for 0-3 hours daily with low levels of physical activity. Kim et al. (2018) identified lifestyle factors such as BMI and alcohol consumption as significant variables in the risk of hypertension, consistent with Okubo et al. (2002), in that smokers tended to have poorer dietary habits, including fruit consumption.

Both studies confirm that public health interventions need to be focused on older adults and smokers. These findings indicate that smoking and changes in lifestyle behavior are the leading drivers of risk for hypertension and that targeting current smokers with lifestyle modification could potentially reduce a large proportion of the prevalence of hypertension. Kim et al., 2018; Okubo et al., 2002 Overall, the findings from the present investigation support the prevailing literature, reinforcing interactions between smoking, hypertension, and the various modifiers.

## Conclusion

This analysis confirms the complex and multifactorial relation between smoking and hypertension, as evidence points out that former smokers have a higher risk of developing hypertension as compared to current smokers, reflecting the smoldering effects of smoking even after cessation. The dose-dependent relation of pack-years to blood pressure and the paradoxical finding of lower blood pressure in current smokers bring into light the complex hemodynamic effects of nicotine. The interaction between smoking and BMI, gender, and other lifestyle factors such as alcohol consumption, sedentary behavior, and poor dietary habits calls for comprehensive public health interventions. Gender disparities in smoking and hypertension-especially the notably higher prevalence among men-indicate the need for sex-specific prevention strategies. Findings indicative of smoking reduction in the context of general lifestyle improvement using dietary improvements and increased physical activity are thus a strategic course of action that will most effectively reduce the risk of hypertension. In the context of the longitudinal impact of smoking cessation on hypertension, future research is warranted as public health policies seek to integrate smoking cessation into a broader set of lifestyle-related interventions.

## Data Availability

The datasets generated and analyzed during the current study are available from the author upon request.

## Acknowledgment

The authors would like to thank all the employees of the beverage company who participated in this study.

## Scientific Responsibility Statement

Authors take the scientific responsibility for the article including the study design, data collection, analysis and interpretation, writing, some of the main line, or all of the preparation and scientific review of the contents and approval of the final version of the article.

## Animal and human rights statement

All procedures performed in studies involving human participants were by the ethical standards of the institutional and/or national research committee and with the 1964 Helsinki Declaration and its later amendments or comparable ethical standards. No animal or human studies were carried out by the authors of this article.

## References

1. Abdullahi A, et al. Smoking as a risk factor for chronic non-specific low back pain: a systematic review and meta-analysis. Chiropr Man Therap. 2022;30(1):1–11.

2. Action on Smoking and Health. Smoking and Cardiovascular Disease [Internet]. Action on Smoking and Health; 2016 [cited 2024 Feb 4]. Available from: https://ash.org.uk/information-and-resources/fact-sheets/smoking-and-cardiovascular-disease/

3. Ahmed T, et al. Prevalence of hypertension and its associated factors among the elderly population in rural Bangladesh. Heliyon. 2019;5(10):e02619.

4. Ain QU, Regmi K. The effects of smoking in developing hypertension in Pakistan: a systematic review. South East Asia J Public Health. 2015;5(1):4–11.

5. Akpa OM, Okekunle AP, Asowata JO, Adedokun B. Passive smoking exposure and the risk of hypertension among non-smoking adults: the 2015–2016 NHANES data. Clin Hypertens. 2021;27:1–12.

6. Alberg AJ, et al. The association of cigarette smoking with circulating concentrations of proinflammatory biomarkers. Circulation. 2014;129(6):537–45.

7. Bhagyalaxmi A, et al. Prevalence and pattern of smoking among adult population in rural Haryana. Indian J Community Med. 2013;38(3):162–6.

8. Bernabe-Ortiz A, Carrillo-Larco RM. Second-hand smoking, hypertension, and cardiovascular risk: findings from Peru. BMC Cardiovasc Disord. 2021;21:1–8.

9. D’Elia L, De Palma D, Rossi G, Strazzullo V, Russo O, Iacone R, et al. Not smoking is associated with lower risk of hypertension: results of the Olivetti Heart Study. Eur J Public Health. 2014;24(2):226–30.

10. Dochi M, Sakata K, Oishi M, Tanaka K, Kobayashi E, Suwazono Y. Smoking as an independent risk factor for hypertension: a 14-year longitudinal study in male Japanese workers. Tohoku J Exp Med. 2009;217(1):37–43.

11. Gao K, Shi X, Wang W. The life-course impact of smoking on hypertension, myocardial infarction, and respiratory diseases. Sci Rep. 2017;7(1):4330.

12. Hering D, Kucharska W, Kara T, Somers VK, Narkiewicz K. Smoking is associated with chronic sympathetic activation in hypertension. Blood Press. 2010;19(3):152–5.

13. Halperin RO, Gaziano MJ, Sesso HD. Smoking and the risk of incident hypertension in middle-aged and older men. Am J Hypertens. 2008;21(2):148–52.

14. Islam MM, et al. Urban-rural differences in prevalence and determinants of undiagnosed hypertension in Bangladesh. Int J Hypertens. 2020;2020.

15. Islam MN, et al. Prevalence and determinants of chronic obstructive pulmonary disease (COPD) in Bangladesh. COPD J Chron Obstruct Pulmon Dis. 2018;15(4):370–8.

16. Islam MS, et al. Prevalence of hypertension in Bangladesh: effect of a socioeconomic risk factor on the difference between rural and urban community. Mymensingh Med J. 2015;24(4):638–46.

17. James WPT, et al. Overweight and obesity (high body mass index). In: Comparative quantification of health risks: global and regional burden of disease attributable to selected major risk factors. Vol. 1. Geneva: World Health Organization; 2014. p. 497–596.

18. John U, Meyer C, Hanke M, Völzke H, Schumann A. Smoking status, obesity and hypertension in a general population sample: a cross-sectional study. J Assoc Physicians. 2006;99(6):407–15.

19. Kaplan RC, et al. Current smoking raises the risk of incident hypertension: Hispanic Community Health Study–Study of Latinos. Am J Hypertens. 2021;34(2):190–7.

20. Khan MM, et al. Prevalence and determinants of hypertension among the elderly population in rural Bangladesh. J Aging Health. 2021;33(5-6):461–9.

21. Khanam MA, et al. Awareness and control of hypertension in Bangladesh: follow-up of a hypertensive cohort. BMJ Open. 2015;5(12):e008034.

22. Li G, et al. The association between smoking and blood pressure in men: a cross-sectional study. BMC Public Health. 2017;17:1–6.

23. Lusno MFD. Association between smoking and hypertension as health burden in Sidoarjo: a case-control study. Int J Appl Biol. 2020;4(2):9–16.

24. Malik MA, et al. Prevalence of hypertension among the adult population in rural Bangladesh: a cross-sectional study. J Public Health Res. 2019;8(2):1666.

25. Malik MA, et al. Bangladesh Cardiovascular Journal: National Heart Foundation of Bangladesh [Internet]. 2020 [cited 2024 Feb 4]. Available from: http://www.bcvj.org/

26. Mendez-Pinto MM, et al. Smoking habits, health disparities, and mental health: a cross-sectional study among university students in Bangladesh. Front Public Health. 2022;10:676915.

27. Morisaki N, et al. Association between smoking and hypertension in pregnancy among Japanese women: a meta-analysis of birth cohort studies in the Japan Birth Cohort Consortium (JBiCC) and JECS. J Epidemiol. 2023;33(10):498–507.

28. Nargis N, et al. Patterns and determinants of tobacco use in Bangladesh from 2009 to 2012: evidence from International Tobacco Control (ITC) study. PLoS One. 2015;10(11):e0141135.

29. Okubo Y, et al. An association between smoking habits and blood pressure in normotensive Japanese men. J Hum Hypertens. 2002;16(2):91–6.

30. Primatesta P, et al. Association between smoking and blood pressure: evidence from the health survey for England. Hypertension. 2001;37(2):187–93.

31. Salam SS, et al. Prevalence and predictors of smoking in Bangladesh: findings from the Global Adult Tobacco Survey. Int J Environ Res Public Health. 2021;19(7):3840.

32. Saladini F, et al. Effects of smoking on central blood pressure and pressure amplification in hypertension of the young. Vasc Med. 2016;21(5):422–8.

33. Sayeed MA, et al. Prevalence of hypertension in Bangladesh: effect of sociodemographic risk factors. J Cardiovasc Dis Res. 2016;7(4):143–8.

34. Segura-Egea JJ, Castellanos-Cosano L, Velasco-Ortega E, Ríos-Santos JV, Llamas-Carreras JM, Machuca G, López-Frías F.

